# Vaccine confidence and timeliness of childhood immunisation by health information source, maternal, socioeconomic, and geographic characteristics in Albania

**DOI:** 10.1101/2021.03.09.21253180

**Authors:** Daniela Mayerová, Kaja Abbas

**Author notes:** Corresponding author: Daniela Mayerová, at 50 rue Geiler, 67 000 Strasbourg, France.

## Abstract

**Background:** Albania is facing decreasing childhood immunisation coverage and timeliness despite a growing economy and universal health insurance. Our aim is to estimate vaccine confidence and timeliness of childhood immunisation by health information source, maternal, socioeconomic, and geographic characteristics in Albania.

**Methods:** We used the 2017-2018 Albania Demographic and Health Survey to analyse vaccine confidence, measured via the proxy of vaccine timeliness, among 2,156 mothers of under-5-year-old children using simple and multivariable logistic regression.

**Results:** 77.9% [74.2, 81.2] of mothers had confidence in vaccines. Immunisation delay was reported by 21.5% [18.3, 25.3] of mothers, but a majority (65.7%) were caused by the infant’s sickness at the time of vaccination, while a minority (8.7%) due to mothers’ concerns about vaccine safety and side effects. Among 1.7% of mothers who ever refused vaccination of their children, the main concerns were about vaccine safety (35.9%) and adverse events (42.7%). Factors associated with lower vaccine confidence were using the Internet/social media as the main health information source compared to other sources (AOR=0.66 [0.47, 0.94], p=0.020), mother’s work outside the home (AOR=0.65 [0.47, 0.91], p=0.013), lack of maternal education (AOR=0.14 [0.03, 0.67], p=0.014), and living in AL02-Qender (AOR=0.38 [0.23, 0.63]) and AL03-Jug regions (AOR=0.36 [0.24, 0.53]) in comparison to AL01-Veri region (p<0.0001).

**Conclusions:** Reinforcement of scientific evidence-based online communication about childhood immunisation and monitoring anti-vaccination movements on the Internet/social media would be beneficial in improving vaccine confidence and timeliness in Albania, together with traditional ways of promoting vaccination by healthcare professionals who enjoy confidence as trusted sources of information.

## Introduction

Immunisation is one of the most important and cost-effective public health interventions [1,2]. However, maintaining a high level of public confidence in vaccines and immunisation programmes and minimizing delay and rejection of vaccination are increasingly challenging worldwide [3–5].

Vaccine confidence refers to the trust in – the effectiveness and safety of vaccines, the reliability of the health service delivery system, the competencies of the health professions, and the motivation of the policymakers in deciding the required vaccines [6]. Vaccine confidence is one of the factors to influence vaccine hesitancy which refers to the delay in acceptance or refusal of vaccines despite availability of vaccine services [6].

Vaccine confidence impacts vaccination demand and is influenced by numerous characteristics and varies widely among countries [4,5,7]. A survey on attitudes towards immunisation in 67 countries revealed that skepticism about vaccine importance and safety is a particularly sensitive issue in Europe, and more serious than in other World Health Organization (WHO) regions [4]. In a related survey conducted in 18 European countries on the attitudes and behaviors among parents regarding their children’s immunisation, 20% of parents delayed vaccination and 12% refused vaccination while 24% of parents identified themselves as somewhat hesitant and 4% as very hesitant [7].

The midterm report of the WHO European Vaccine Action Plan for 2015-2020 reported that decreasing vaccine confidence was contributing to suboptimal immunisation coverage and delays in uptake [8]. The goal of reaching 95% or higher coverage for DTP3 (third dose of diphtheria-tetanus-pertussis) vaccination in 90% of member states was at risk, and elimination of measles and rubella transmission goals for 2015 were not met (ibid). There were 82,596 measles cases during the 2018 outbreak affecting 47 of 53 WHO European region countries, which were 3-times more than in 2017 and 15-times more than in 2016 [9]. Albania, with a population of 2.9 million people, reported 1,469 measles cases during the 2018 outbreak [10].

### Childhood immunisation in Albania

Mandatory vaccination of infants aged 12-23 months in Albania includes BCG (Bacillus Calmette-Guérin) and hepatitis B vaccine at birth, three doses of diphtheria/pertussis/tetanus/hepatitis B/Haemophilus influenzae type b (DPT-HepB-Hib) vaccine (at 2, 4, and 6 months), three doses of polio vaccine (at 2, 4, 6 months), three doses of pneumococcal vaccine (PCV, at 2, 4, 10 months), and one dose of measles-mumps-rubella vaccine (MMR, at 12 months). For infants 24-35 months old, additional mandatory immunisation consists of booster doses of DTP and polio at 2 years [11]. A booster dose of measles vaccine is administered at 5 years [11] and is not considered in this study.

Albania is facing decreasing childhood immunisation coverage despite a growing economy (post-communist upper-middle-income country), universal health insurance [12], rising expenditure in health services [11], and free of charge childhood immunisation [13]. The proportion of zero dose children (children who never received any vaccines) was below 1% in 2017-18 [12] as it was in 2008-2009 [14]. However, all basic vaccination coverage in 12-23 months old infants declined from 94% in 2008-2009 to 75% in 2017-2018 (BCG, three doses of DPT-HepB-Hib, three doses of polio vaccine, and one dose of measles-containing vaccine) [12]. While in 2017-2018, 75.0% and 87.9% of infants 12-23 months old and 24-35 months old respectively received all basic vaccinations, rates of all age-appropriate vaccines were 67.2% in infants 12-23 months old and 74.5% in children 24-35 months old [12]. These differences indicate a substantial immunisation delay.

### Health information source, maternal, child, socioeconomic, and geographic determinants of vaccine confidence

The Internet is a valuable source of information related to vaccines and health in general but also spread substantial misinformation regarding immunisation [15–17]. Though most documents expected to be retrieved by a person seeking information on vaccines are pro-vaccination [17–21], research indicates a relationship between the parental online search for childhood vaccines and more negative attitudes towards immunisation compared to parents with other sources of health-related information [22–25].

A recent systematic review described child gender, maternal education and age, socioeconomic status, urban/rural residence, household size, ethnicity, birth setting, child age, birth order, maternal occupation, marital status, paternal education, distance to clinic, antenatal care visits, maternal and parental occupation, birth year, religion, region, and paternal age as the most commonly tested predictors of childhood immunisation timeliness in low- and middle-income countries [26]. Though Albanian children’s immunisation pattern is similar to that in high-income countries – achieved high vaccination uptake, followed by a decline over the last decade – social determinants of health persist in Europe [27,28], and most characteristics identified by this systematic review are considered relevant in Albania.

### Study objective

The primary focus of this study is on vaccine confidence, and timeliness of childhood immunisation is used as a proxy measure of vaccine confidence among mothers in Albania. Timeliness refers to adherence to immunisation schedules, that is no delay or refusal of a vaccine. We analysed the 2017-2018 Albania Demographic and Health Survey (DHS) [29] to assess the vaccine confidence and timeliness of childhood immunisation by health information source, maternal, child, socioeconomic, and geographic characteristics in Albania.

## Methods

### Survey data

Demographic and Health Surveys have been conducted in over 90 low- and middle-income countries [29]. These surveys collect data on fertility and family planning, maternal and child health, knowledge and attitudes toward HIV/AIDS and other sexually transmitted infections, and prevalence of non-communicable diseases.

We used the 2017-2018 Albania Demographic and Health Survey to estimate vaccine confidence and timeliness of childhood immunisation [12]. The survey interview was completed by 10,860 women aged 15-49 years (93% of those eligible), and data were collected for 2,762 children born to these women within the last five years (before survey interview). We excluded data for 435 children due to missing data on vaccination delay and refusal, and we used sample weighted data for 2,156 mother-child pairs in our analysis, as shown in Figure 1.

**Figure 1:**
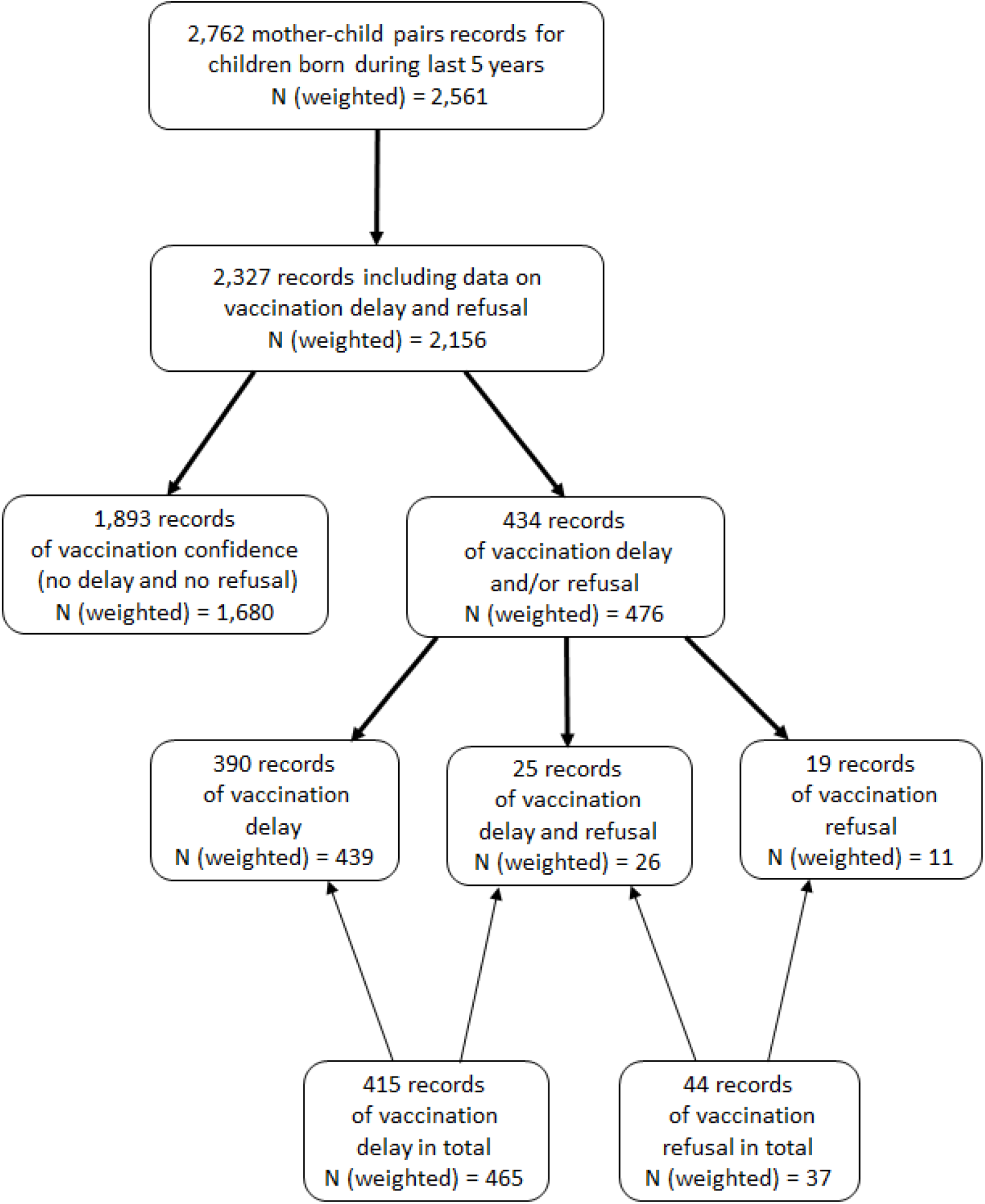
Flow-chart illustrating subpopulations for vaccine confidence from the Demographic and Health survey. Vaccine confidence was measured through the proxy measure of vaccine timeliness among the different subpopulations in the 2017-2018 Albania Demographic and Health Survey – vaccination on-time with no delay or refusal, vaccination delay, vaccination delay and refusal, and vaccination refusal.

### Vaccine confidence analysis

The outcome variable vaccine confidence was defined as no delaying and no refusing childhood immunisation, that is answers “no” to questions “Have you ever postponed or delayed having your child (one of your children) vaccinated?” and “Have you ever chosen not to have your child (one of your children) vaccinated?”. The independent variables of interest were based on health information source, maternal, child, socioeconomic, and geographic characteristics. Health information source characteristics include Internet/social media as main source of health information and trusted sources of information on vaccines; maternal characteristics include age of childbirth, education, marital status, and household head status; child characteristics include gender and birth order; socioeconomic characteristics include mother’s work status, partner’s work status, partner’s education, household wealth, ethnicity, and religion; and geographic characteristics include area of residence (urban/rural) and prefecture.

We conducted simple logistic regression to estimate crude odds ratios and assess vaccine confidence disaggregated by the above mentioned characteristics using all available observations (N weighted=2,156). Then we excluded 48 records (2%) with missing data (22 on Internet/social media, and 26 for each of the partner’s education and work) and followed up with multivariable logistic regression in a complete dataset (N weighted=2,113) to estimate the association of selected independent variables with the dependent outcome variable of vaccine confidence, after controlling for other background characteristics. Variables selected for the multivariable model were those associated with vaccine confidence in the univariate analysis, defined as with p-values <0.05 and/or having a strong effect on vaccine confidence with odds ratio – OR ≤ 0.5 or OR ≥ 1.5. Finally, we investigated potential for interaction by including an interaction term between Internet/social media and mother’s education, and between mother’s work and partner’s work.

### Ethics approval and reproducible analysis

This study was approved by the ethics committee (Ref 21362) of the London School of Hygiene & Tropical Medicine. The 2017-2018 Albania DHS dataset is accessible upon registration on the DHS website [29]. The survey analysis was conducted by taking into account stratification, cluster sampling, and sampling weights, using Stata statistical software [30], and the code is accessible at https://github.com/d-mayerova/Vaccine_confidence_Albania.

## Results

### Characteristics of the study population

Table 1 includes the characteristics of the study population. 30.0% of participating mothers reported use of the Internet as the main source of information about health; other main sources were healthcare providers (42.1%), TV (24.7%), friends/relatives (2.3%), schools (0.5%), newspapers (0.4%), radio (0.1%) and other (0.2%). Most women considered healthcare professionals the most trusted source of information on vaccines. More than one-quarter of mothers completed higher education, while 1.2% did not receive any training. Most respondents were Albanian, Muslim, married/living with a partner, and not working outside the home during 12 months preceding the survey. Children born during the last five years before the survey were 50.4% males and 43.5% were first-born. Most households were headed by men at 84.6%. The proportion of partners of the participating women who did not receive any education was similar to women at 1.4%. The majority of partners at least partially worked during the 12 months preceding the survey at 81.2%. The study population was slightly more urban (56.3%) than rural.

**Table 1:**
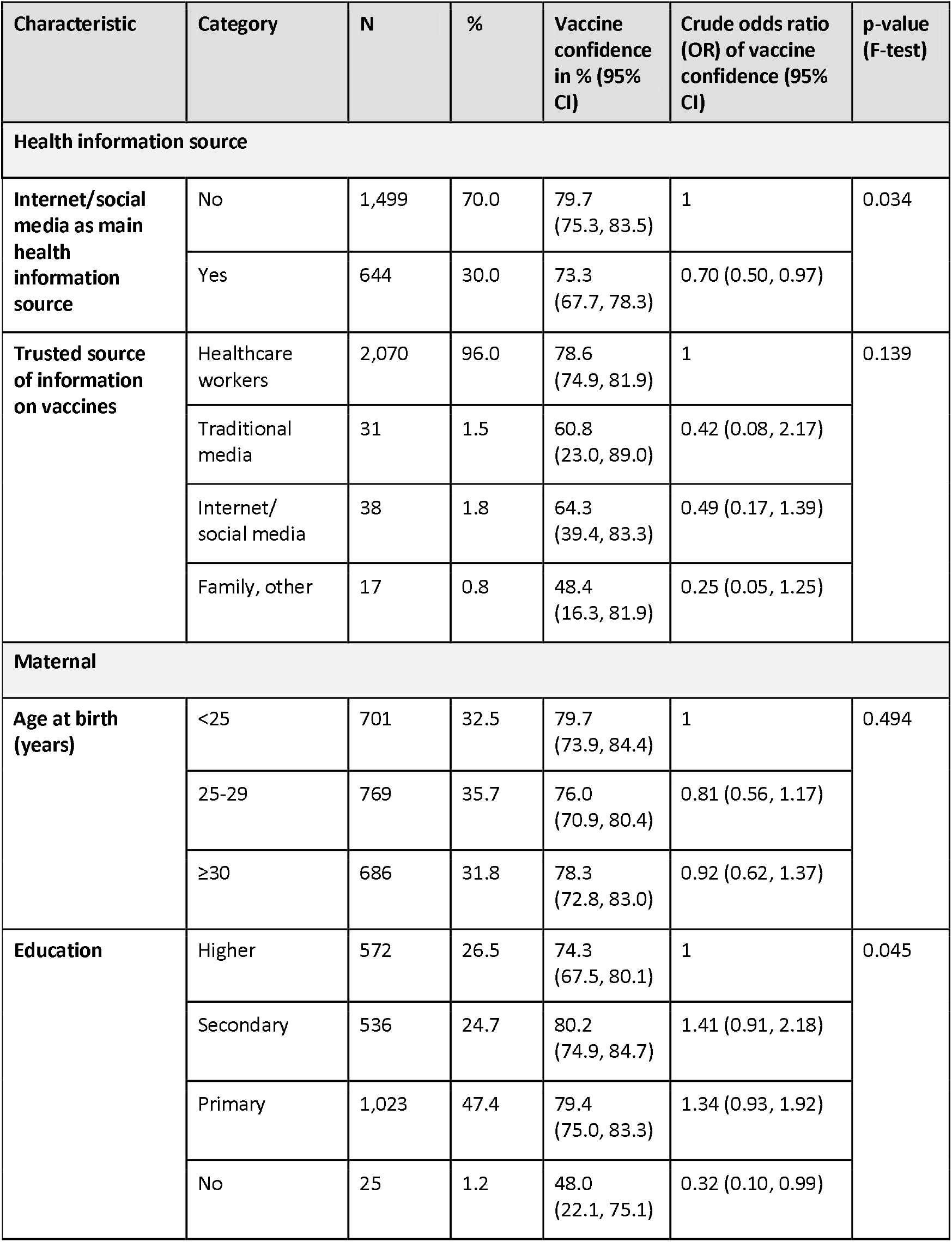
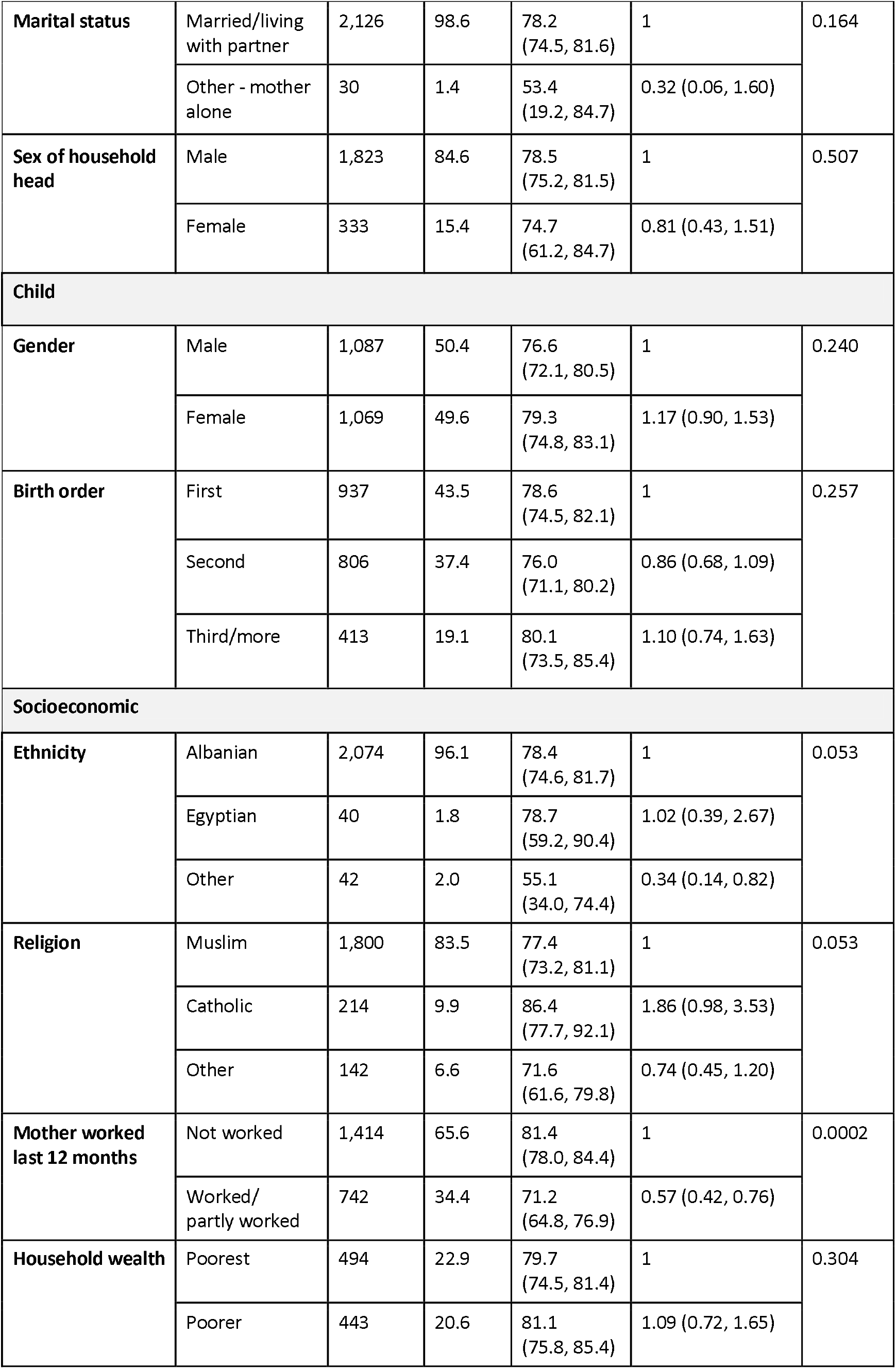
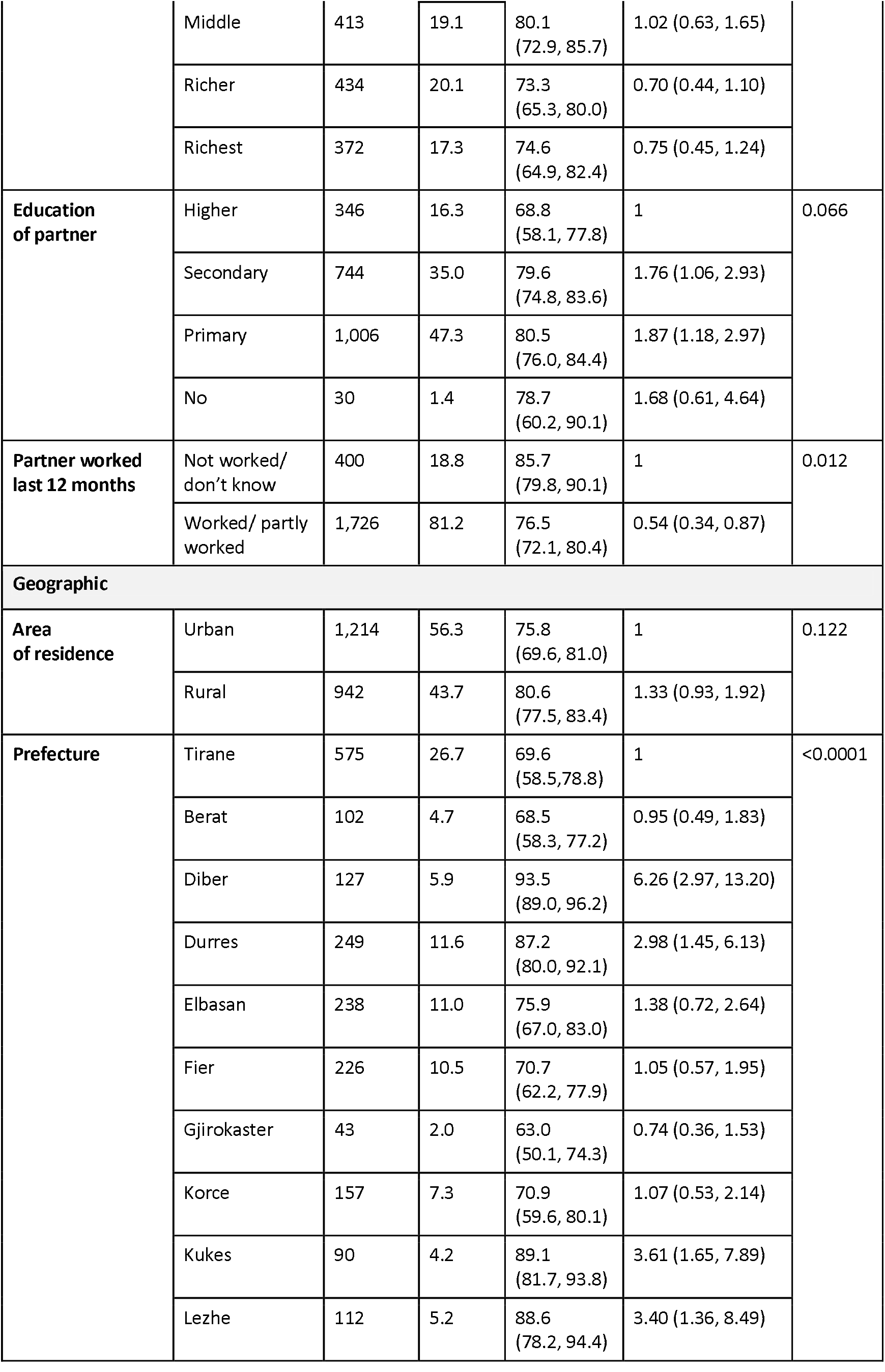
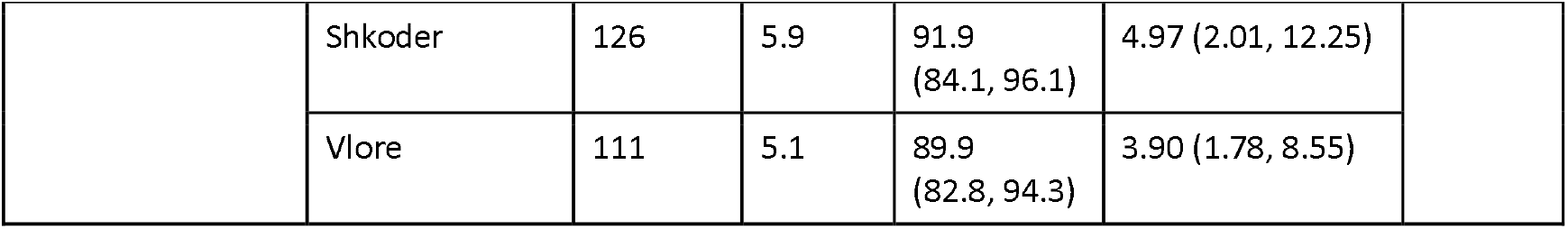
Vaccine confidence among mothers of under-5-year-old children in childhood immunisation disaggregated by health information source, socioeconomic, geographic, maternal, and child characteristics in Albania. Crude odds ratios for vaccine confidence by these characteristics were estimated via simple logistic regression.

| Characteristic | Category | N | % | Vaccine confidence in % (95% CI) | Crude odds ratio (OR) of vaccine confidence (95% CI) | p-value (F-test) |
| --- | --- | --- | --- | --- | --- | --- |
| Health information source |  |  |  |  |  |  |
| Internet/social media as main health information source | No | 1,499 | 70.0 | 79.7 (75.3, 83.5) | 1 | 0.034 |
|  | Yes | 644 | 30.0 | 73.3 (67.7, 78.3) | 0.70 (0.50, 0.97) |  |
| Trusted source of information on vaccines | Healthcare workers | 2,070 | 96.0 | 78.6 (74.9, 81.9) | 1 | 0.139 |
|  | Traditional media | 31 | 1.5 | 60.8 (23.0, 89.0) | 0.42 (0.08, 2.17) |  |
|  | Internet/ social media | 38 | 1.8 | 64.3 (39.4, 83.3) | 0.49 (0.17, 1.39) |  |
|  | Family, other | 17 | 0.8 | 48.4 (16.3, 81.9) | 0.25 (0.05, 1.25) |  |
| Maternal |  |  |  |  |  |  |
| Age at birth (years) | <25 | 701 | 32.5 | 79.7 (73.9, 84.4) | 1 | 0.494 |
|  | 25-29 | 769 | 35.7 | 76.0 (70.9, 80.4) | 0.81 (0.56, 1.17) |  |
|  | ≥30 | 686 | 31.8 | 78.3 (72.8, 83.0) | 0.92 (0.62, 1.37) |  |
| Education | Higher | 572 | 26.5 | 74.3 (67.5, 80.1) | 1 | 0.045 |
|  | Secondary | 536 | 24.7 | 80.2 (74.9, 84.7) | 1.41 (0.91, 2.18) |  |
|  | Primary | 1,023 | 47.4 | 79.4 (75.0, 83.3) | 1.34 (0.93, 1.92) |  |
|  | No | 25 | 1.2 | 48.0 (22.1, 75.1) | 0.32 (0.10, 0.99) |  |
| Marital status | Married/living with partner | 2,126 | 98.6 | 78.2<br>(74.5, 81.6) | 1 | 0.164 |
|  | Other - mother alone | 30 | 1.4 | 53.4<br>(19.2, 84.7) | 0.32 (0.06, 1.60) |  |
| Sex of household head | Male | 1,823 | 84.6 | 78.5<br>(75.2, 81.5) | 1 | 0.507 |
|  | Female | 333 | 15.4 | 74.7<br>(61.2, 84.7) | 0.81 (0.43, 1.51) |  |
| Child |  |  |  |  |  |  |
| Gender | Male | 1,087 | 50.4 | 76.6<br>(72.1, 80.5) | 1 | 0.240 |
|  | Female | 1,069 | 49.6 | 79.3<br>(74.8, 83.1) | 1.17 (0.90, 1.53) |  |
| Birth order | First | 937 | 43.5 | 78.6<br>(74.5, 82.1) | 1 | 0.257 |
|  | Second | 806 | 37.4 | 76.0<br>(71.1, 80.2) | 0.86 (0.68, 1.09) |  |
|  | Third/more | 413 | 19.1 | 80.1<br>(73.5, 85.4) | 1.10 (0.74, 1.63) |  |
| Socioeconomic |  |  |  |  |  |  |
| Ethnicity | Albanian | 2,074 | 96.1 | 78.4<br>(74.6, 81.7) | 1 | 0.053 |
|  | Egyptian | 40 | 1.8 | 78.7<br>(59.2, 90.4) | 1.02 (0.39, 2.67) |  |
|  | Other | 42 | 2.0 | 55.1<br>(34.0, 74.4) | 0.34 (0.14, 0.82) |  |
| Religion | Muslim | 1,800 | 83.5 | 77.4<br>(73.2, 81.1) | 1 | 0.053 |
|  | Catholic | 214 | 9.9 | 86.4<br>(77.7, 92.1) | 1.86 (0.98, 3.53) |  |
|  | Other | 142 | 6.6 | 71.6<br>(61.6, 79.8) | 0.74 (0.45, 1.20) |  |
| Mother worked last 12 months | Not worked | 1,414 | 65.6 | 81.4<br>(78.0, 84.4) | 1 | 0.0002 |
|  | Worked/<br>partly worked | 742 | 34.4 | 71.2<br>(64.8, 76.9) | 0.57 (0.42, 0.76) |  |
| Household wealth | Poorest | 494 | 22.9 | 79.7<br>(74.5, 81.4) | 1 | 0.304 |
|  | Poorer | 443 | 20.6 | 81.1<br>(75.8, 85.4) | 1.09 (0.72, 1.65) |  |
|  | Middle | 413 | 19.1 | 80.1<br>(72.9, 85.7) | 1.02 (0.63, 1.65) |  |
|  | Richer | 434 | 20.1 | 73.3<br>(65.3, 80.0) | 0.70 (0.44, 1.10) |  |
|  | Richest | 372 | 17.3 | 74.6<br>(64.9, 82.4) | 0.75 (0.45, 1.24) |  |
| Education of partner | Higher | 346 | 16.3 | 68.8<br>(58.1, 77.8) | 1 | 0.066 |
|  | Secondary | 744 | 35.0 | 79.6<br>(74.8, 83.6) | 1.76 (1.06, 2.93) |  |
|  | Primary | 1,006 | 47.3 | 80.5<br>(76.0, 84.4) | 1.87 (1.18, 2.97) |  |
|  | No | 30 | 1.4 | 78.7<br>(60.2, 90.1) | 1.68 (0.61, 4.64) |  |
| Partner worked last 12 months | Not worked/<br>don't know | 400 | 18.8 | 85.7<br>(79.8, 90.1) | 1 | 0.012 |
|  | Worked/ partly<br>worked | 1,726 | 81.2 | 76.5<br>(72.1, 80.4) | 0.54 (0.34, 0.87) |  |
| Geographic |  |  |  |  |  |  |
| Area of residence | Urban | 1,214 | 56.3 | 75.8<br>(69.6, 81.0) | 1 | 0.122 |
|  | Rural | 942 | 43.7 | 80.6<br>(77.5, 83.4) | 1.33 (0.93, 1.92) |  |
| Prefecture | Tirane | 575 | 26.7 | 69.6<br>(58.5,78.8) | 1 | <0.0001 |
|  | Berat | 102 | 4.7 | 68.5<br>(58.3, 77.2) | 0.95 (0.49, 1.83) |  |
|  | Diber | 127 | 5.9 | 93.5<br>(89.0, 96.2) | 6.26 (2.97, 13.20) |  |
|  | Durres | 249 | 11.6 | 87.2<br>(80.0, 92.1) | 2.98 (1.45, 6.13) |  |
|  | Elbasan | 238 | 11.0 | 75.9<br>(67.0, 83.0) | 1.38 (0.72, 2.64) |  |
|  | Fier | 226 | 10.5 | 70.7<br>(62.2, 77.9) | 1.05 (0.57, 1.95) |  |
|  | Gjirokaster | 43 | 2.0 | 63.0<br>(50.1, 74.3) | 0.74 (0.36, 1.53) |  |
|  | Korce | 157 | 7.3 | 70.9<br>(59.6, 80.1) | 1.07 (0.53, 2.14) |  |
|  | Kukes | 90 | 4.2 | 89.1<br>(81.7, 93.8) | 3.61 (1.65, 7.89) |  |
|  | Lezhe | 112 | 5.2 | 88.6<br>(78.2, 94.4) | 3.40 (1.36, 8.49) |  |
|  | Shkoder | 126 | 5.9 | 91.9<br>(84.1, 96.1) | 4.97 (2.01, 12.25) |  |
|  | Vlore | 111 | 5.1 | 89.9<br>(82.8, 94.3) | 3.90 (1.78, 8.55) |  |

### Childhood immunisation delay and refusal

77.9% (74.2, 81.2) of mothers never delayed and never refused childhood immunisation, approximating the overall level of confidence in childhood vaccines. 21.5% (18.3, 25.3) of mothers reported immunisation delay, and 1.7% (1.0, 3.1) ever chose not to have their child/children vaccinated.

Among mothers who delayed childhood vaccination, nearly two-thirds of them attributed the delay to the sickness of their child at the time of vaccination. Around 10% of the mothers attributed the delay due to lack of time while another around 10% of mothers indicated other reasons than those listed in the questionnaire (see Table 2). Vaccine safety and side effects were less common (around 8% combined) stated reasons for delaying vaccination, and a small proportion (6%) of mothers had no particular reason for immunisation delay. Among mothers who refused childhood immunisation, the main reasons were attributed to concerns about vaccine safety and side effects (see Table 3).

**Table 2:** Reasons for delaying childhood immunisation. Reasons stated by mothers of under-5-year-old children to delay childhood immunisation in Albania.

| Reason for delaying childhood vaccination | N | Proportion in % (95% CI) |
| --- | --- | --- |
| Doubts about vaccine safety | 14 | 2.9 (1.4, 6.2) |
| Concerns about side effects | 24 | 5.2 (2.2, 11.9) |
| Child was sick at the time of vaccination | 305 | 65.7 (57.6, 73.0) |
| Did not have time/too busy | 43 | 9.3 (5.5, 15.4) |
| No particular reason | 28 | 6.0 (3.6, 9.9) |
| Other | 50 | 10.8 (7.3, 15.8) |

**Table 3:** Reasons for refusing childhood immunisation. Reasons stated by mothers of under-5-year-old children to refuse childhood immunisation in Albania.

| Reason | N | Proportion in % (95% CI) |
| --- | --- | --- |
| Doubts about vaccine safety | 13 | 35.9 (15.2, 63.8) |
| Concerns about side effects | 16 | 42.7 (15.4, 75.4) |
| Child was sick at the time of vaccination | 1 | 3.8 (1.0, 13.0) |
| Medical contraindication | 0 | 0 |
| Vaccine more harmful than disease | 2 | 5.1 (1.0, 21.4) |
| Religious conviction | 0 | 0 |
| Other | 5 | 12.5 (3.7, 34.7) |

### Vaccine confidence

Vaccine confidence among mothers of under-5-year-old children in childhood immunisation disaggregated by health information source, maternal, child, socioeconomic, and geographic characteristics in Albania are illustrated in Figure 2 and Table 1. Table 1 also includes the crude odds ratios for vaccine confidence by these characteristics that were estimated via simple logistic regression. Mothers using the Internet/social media as the main health information source were less likely confident in vaccination than mothers with other main health information sources, as were mothers whose most trusted source of information on vaccines was other than healthcare professionals. Mothers of ethnic minorities like Roma (the “other” category) were less likely confident in childhood immunisation than Albanian, and less likely confident were also mothers living alone compared to those married/living with a partner. Catholic mothers were more likely confident in vaccination than women of other religions. Vaccine confidence was lower among mothers with higher education in comparison to mothers with primary education, and a similar pattern is observed with lower vaccine confidence among mothers whose partners have higher education in comparison to partners with primary education. Working mothers and those whose partners work had lower vaccine confidence in comparison to those not working/with partners who do not work. Mothers belonging to the top two household wealth quintiles had relatively lower confidence than mothers in the remaining lower wealth quintiles. Vaccine confidence was similar across different maternal ages at birth, similar across both male- and female-headed households, and also similar by child characteristics of gender and birth order. While vaccine confidence was similar by urban and rural areas of residence, it was relatively high in the prefectures of AL01-Veri region (all > 80%, Shkoder and Diber > 90%), followed by prefectures (Elbsan of AL02-Qender region (71 - 80%), and relatively low in the prefectures of AL03-Jug region (< 70%) except for Vlore (see Figure 3).

**Figure 2:**
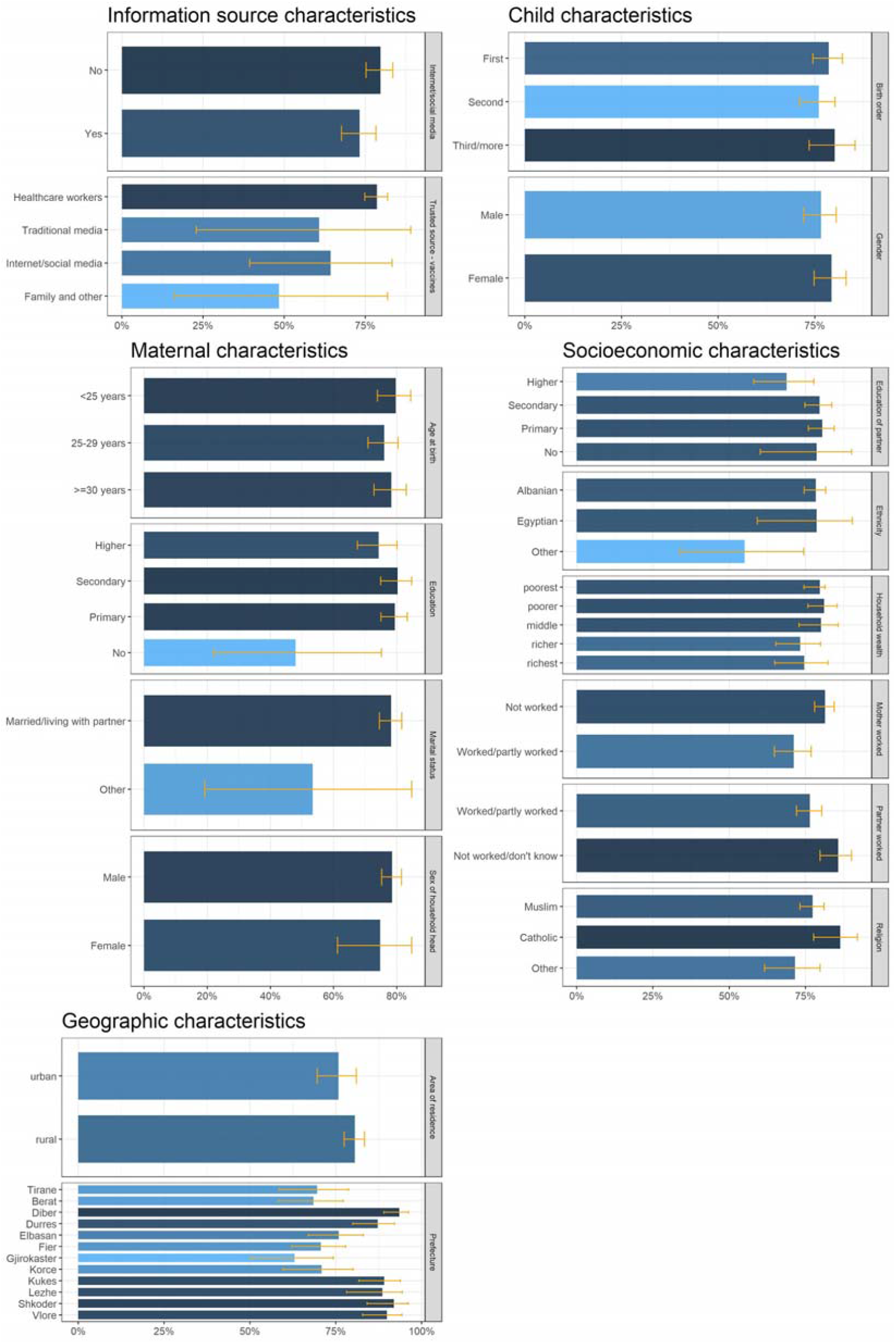
Vaccine confidence among mothers in Albania. Vaccine confidence among mothers of under-5-year-old children in childhood immunisation disaggregated by health information source, child, maternal, socioeconomic, and geographic characteristics in Albania.

**Figure 3:**
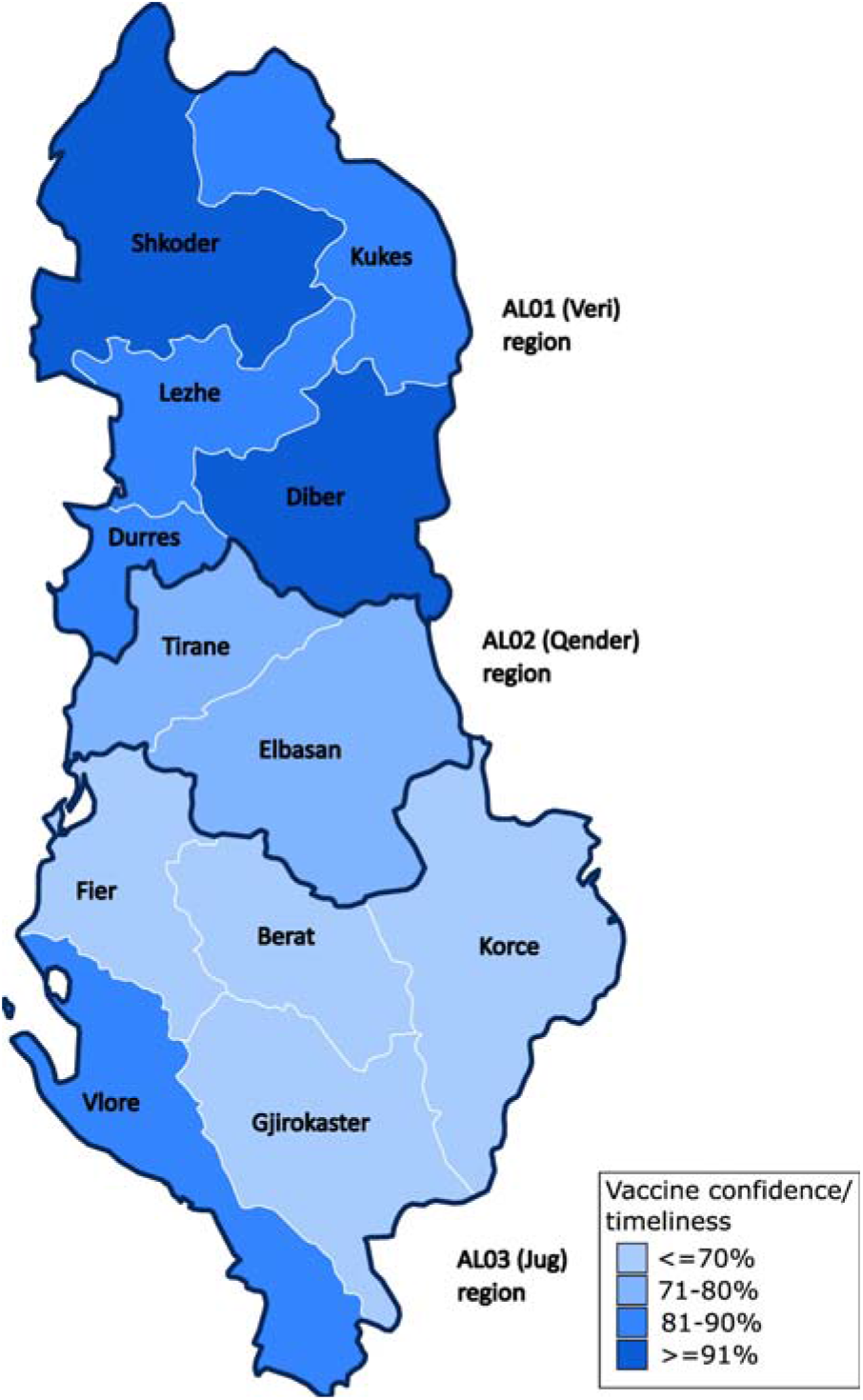
Vaccine confidence among mothers by prefectures and regions in Albania. Vaccine confidence among mothers of under-5-year-old children in childhood immunisation is higher in the AL01-Veri region, followed by AL02-Qender region and the AL03-Jug region.

Variables considered for the multivariable model included Internet/social media as main health information source, trusted source of information on vaccines, maternal education, education of partner, mother’s work, partner’s work, ethnicity, religion, and region. Marital status could not be analysed because there were no respondents in the category “other” following the exclusion of records with missing data on the partner’s education and work. Trusted source of information on vaccines is on the causal pathway between the Internet/social media and vaccine confidence, and was therefore excluded. Ethnicity and religion were excluded on the basis of multicollinearity. To avoid data sparsity, prefectures according to Eurostat NUTS3 division were grouped into NUTS2 regions [31] with minimal loss of precision (see Figure 3).

Vaccine confidence among mothers of under-5-year-old children in childhood immunisation associated with health information source (Internet/social media), maternal (education), socioeconomic (mother’s work, partner’s work, education of partner), and geographic (region) characteristics are illustrated in Figure 4 and Table 4. Adjusted odds ratios for vaccine confidence by these characteristics were estimated via multivariable logistic regression. After controlling for other characteristics, mothers whose main health information source was the Internet/social media had 34% lower odds of confidence in childhood immunisation than mothers with other main information sources, AOR=0.66 (0.47, 0.94), p=0.020. Compared to non-working mothers, those who worked had 35% lower odds of vaccine confidence, AOR=0.65 (0.47, 0.91), p=0.013. Mothers residing in AL02-Qender and AL03-Jug regions had 62% and 64% lower odds of confidence in childhood vaccination, respectively, in comparison to women living in AL01-Veri region; adjusted OR=0.38 (0.23, 0.63) and 0.36 (0.24, 0.53), p<0.0001. Education levels of partner and work status of partner were not associated with confidence in childhood immunisation. Mothers without education had 86% lower odds of vaccine confidence compared to those who completed higher education, AOR=0.14 (0.03, 0.67), p=0.014; there was some weak association (p=0.060) of decreasing vaccine confidence with decreasing education level. There was insufficient evidence for interaction between the Internet/social media and mother’s education (p=0.358), and between mother’s work and partner’s work (p=0.112).

**Table 4:**
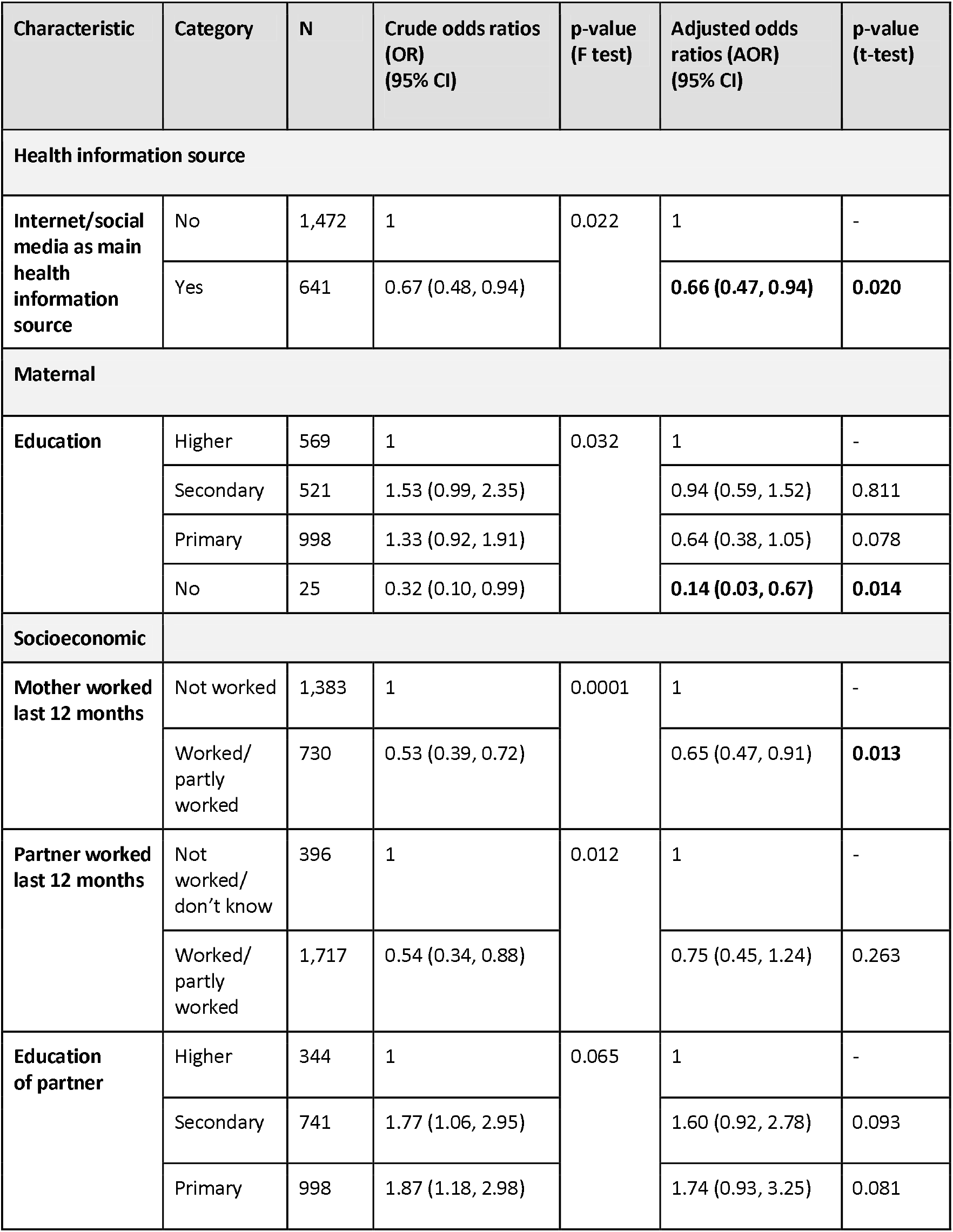
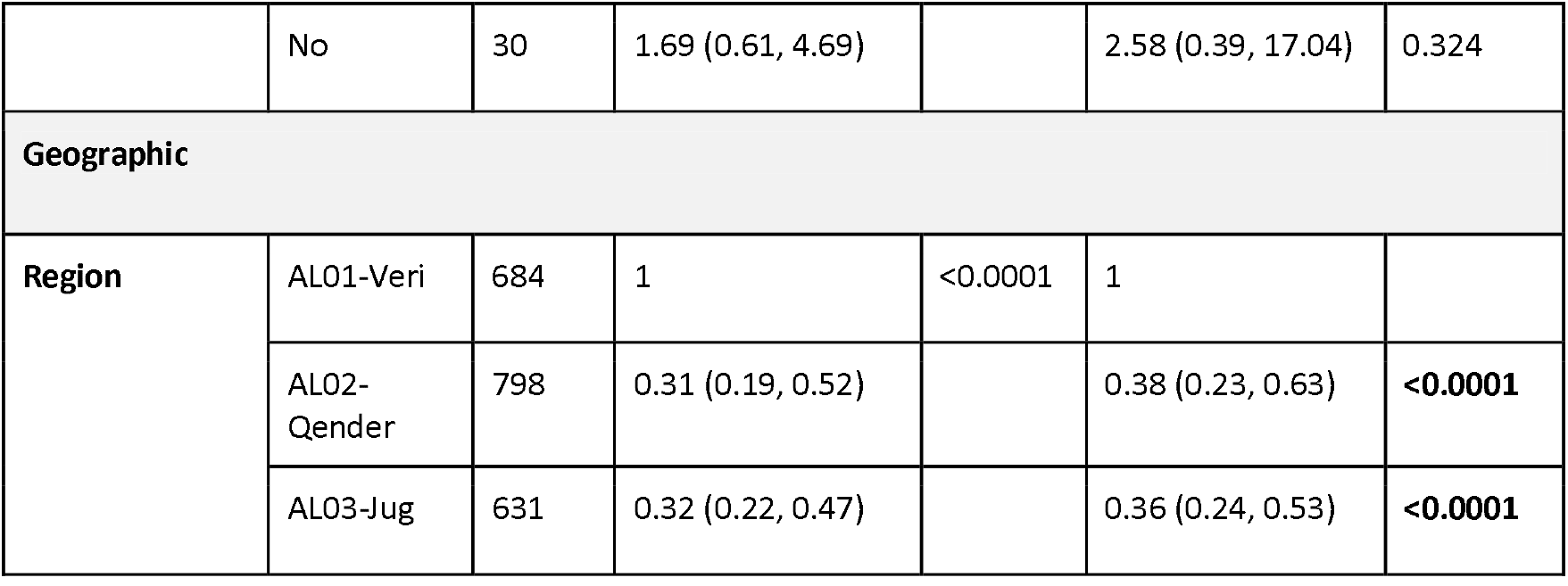
Vaccine confidence among mothers associated with health information source, maternal, socioeconomic, and geographic characteristics in Albania. Vaccine confidence among mothers of under-5-year-old children in childhood immunisation associated with health information source (Internet/social media), maternal (education), socioeconomic (mother’s work, partner’s work, education of partner), and geographic (region) characteristics. Adjusted odds ratios for vaccine confidence by these characteristics were estimated via multivariable logistic regression.

| Characteristic | Category | N | Crude odds ratios (OR) (95% CI) | p-value (F test) | Adjusted odds ratios (AOR) (95% CI) | p-value (t-test) |
| --- | --- | --- | --- | --- | --- | --- |
| Health information source |  |  |  |  |  |  |
| Internet/social media as main health information source | No | 1,472 | 1 | 0.022 | 1 | - |
|  | Yes | 641 | 0.67 (0.48, 0.94) |  | **0.66 (0.47, 0.94)** | **0.020** |
| Maternal |  |  |  |  |  |  |
| Education | Higher | 569 | 1 | 0.032 | 1 | - |
|  | Secondary | 521 | 1.53 (0.99, 2.35) |  | 0.94 (0.59, 1.52) | 0.811 |
|  | Primary | 998 | 1.33 (0.92, 1.91) |  | 0.64 (0.38, 1.05) | 0.078 |
|  | No | 25 | 0.32 (0.10, 0.99) |  | **0.14 (0.03, 0.67)** | **0.014** |
| Socioeconomic |  |  |  |  |  |  |
| Mother worked last 12 months | Not worked | 1,383 | 1 | 0.0001 | 1 | - |
|  | Worked/ partly worked | 730 | 0.53 (0.39, 0.72) |  | 0.65 (0.47, 0.91) | **0.013** |
| Partner worked last 12 months | Not worked/ don't know | 396 | 1 | 0.012 | 1 | - |
|  | Worked/ partly worked | 1,717 | 0.54 (0.34, 0.88) |  | 0.75 (0.45, 1.24) | 0.263 |
| Education of partner | Higher | 344 | 1 | 0.065 | 1 | - |
|  | Secondary | 741 | 1.77 (1.06, 2.95) |  | 1.60 (0.92, 2.78) | 0.093 |
|  | Primary | 998 | 1.87 (1.18, 2.98) |  | 1.74 (0.93, 3.25) | 0.081 |
|  | No | 30 | 1.69 (0.61, 4.69) |  | 2.58 (0.39, 17.04) | 0.324 |
| **Geographic** |  |  |  |  |  |  |
| **Region** | AL01-Veri | 684 | 1 | <0.0001 | 1 |  |
|  | AL02-Qender | 798 | 0.31 (0.19, 0.52) |  | 0.38 (0.23, 0.63) | **<0.0001** |
|  | AL03-Jug | 631 | 0.32 (0.22, 0.47) |  | 0.36 (0.24, 0.53) | **<0.0001** |

**Figure 4:**
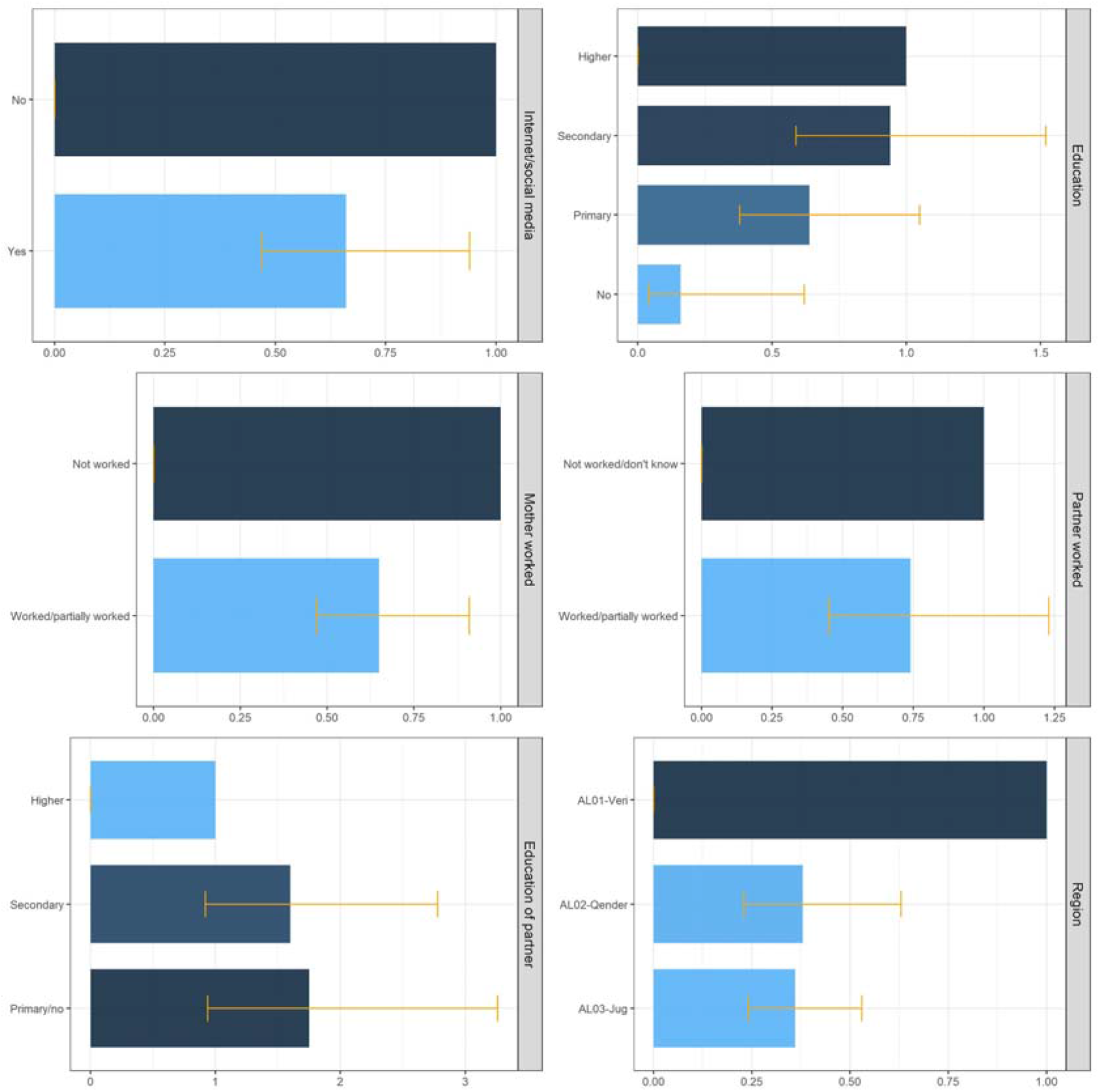
Vaccine confidence among mothers associated with health information source, maternal, socioeconomic, and geographic characteristics in Albania. Vaccine confidence among mothers of under-5-year-old children in childhood immunisation associated with health information source (Internet/social media), maternal (education), socioeconomic (mother’s work, partner’s work), and geographic (region) characteristics in Albania.

## Discussion

To the best of our knowledge, this is the first study to assess vaccine confidence and timeliness of childhood immunisation by health information source, maternal, child, socioeconomic, and geographic characteristics. We examined most of the sociodemographic characteristics identified as risk factors for inequity in vaccination timeliness by systematic reviews [26,32] in a nationally representative sample of mothers with under-5-year-old children.

### Main findings and comparison with other studies

More than three-quarters of mothers were confident in childhood vaccines. Approximately one in five mothers reported immunisation delay, and one in sixty ever refused a vaccine. Most of the delayed vaccines were due to the sickness of the child at the time of vaccination; other reasons included lack of time or doubts about vaccine safety and side effects. Concerns about vaccine safety and side effects were the most frequently reported reason for vaccine refusal. Lower vaccine confidence was associated with having the Internet/social media as the main health information source in comparison to other sources, mother’s work outside the home compared to being in a household, and living in AL02-Qender and AL03-Jug regions in contrast to AL01-Veri. Mothers without education had lower odds of vaccine confidence compared to those who completed higher education.

With regard to other studies, a survey about immunisation concerns among Albanian parents reported higher vaccination timeliness, a lower rate of delay, and a higher vaccine refusal rate [13]. Immunisation delay in our study is similar to the situation in other countries of the Balcan peninsula, Croatia [33], and the neighbouring country of Greece for traditional vaccines (timeliness at 12 and 24 months [34]. Our estimate for childhood immunisation delay of 21.5% from the 2017-2018 Albania DHS survey is similar to the survey estimates published in 2020 for average childhood immunisation delay from 18 European countries of 20% [7]. However, the European survey excluded delays due to child sickness and medical contraindications while child sickness was cited as the main reason for immunisation delay by nearly two-thirds of mothers in the 2017-2018 Albania DHS survey. This suggests that childhood immunisation delay is relatively lower in Albania and thereby higher levels of vaccine confidence than the average of the 18 surveyed European countries. We found a considerably lower vaccine rejection rate than the European [7] and Croatian [33] surveys. Data obtained from immunisation registries for a sub-sample of children of the interviewed mothers – 925 children up to 35 months old – revealed a higher immunisation delay of 29% and refusal of 19% (missing one or more basic vaccinations) [12]. However, the registry-based data did not include older 3-year and 4-year old children whose immunisation might be more compliant with schedules compared to infants up to 35 months old, supporting the DHS finding of increasing vaccination delay/refusal rates. Also, various other reasons may contribute to this difference, like chance, poor memory recall or social desirability bias leading to under-reporting of alternative immunisation schedules, or disproportionally high non-response to questions on vaccine timeliness among mothers who delay/refuse childhood vaccination.

The time-frame for vaccination delay was not defined in the questionnaire. This may result in over-reporting of the delay compared to studies where vaccine timeliness was obtained from immunisation records and checked against a predefined time-frame. However, the opposite effect is also possible, depending on the chosen time-frame (in the absence of an internationally agreed definition of vaccine timeliness [26]).

With regard to reasons for childhood immunisation delay/refusal, our finding that the majority of delays may be rather consequences of adverse circumstances (sickness of the child, mother too busy) than an intention is consistent with the literature [7,35,36]. The survey examining vaccine concerns in Albania [13] did not examine delay and refusal separately, but concerns about safety and adverse effects were the most frequently reported. Overall, doubts about vaccine safety belong to the most critical determinants of vaccine confidence in Europe [4,5]. Further, around 10% of women who delayed or refused immunisation had other reasons than those specified in the questionnaire. Two studies among Albanian caregivers identified additional concerns that might have possibly contributed to the “other” reason category - too many vaccines given to infants [13,37], distrust in vaccines/effectiveness of vaccines provided by the state, and lack of information about vaccination [13].

Lower confidence in childhood immunisation (higher hesitancy) observed in this study among mothers who use mainly the Internet/social media for health information was described as well in France [22], while studies from Poland [38] and the United States [39] found no association between the Internet as vaccine information source and alternative childhood vaccination schedules. Also other studies indicated an adverse impact of the Internet on childhood immunisation, though not through the same relationship Internet versus vaccine timeliness/confidence but with regard to the role of the Internet in vaccination decisions [24] or perception of vaccine safety/effectiveness [40].

Concerning inequities by sociodemographic factors, the negative impact of mother’s work outside the home on childhood immunisation timeliness/confidence is consistent with findings published in a recent systematic review [32] and analysis of DHS surveys from 96 countries [41]. Mothers working outside the home may encounter more difficulties when planning immunisation visits than mothers in households, however, lack of time was reported only by less than one-tenth of the respondents who delayed childhood vaccination. The lower vaccine confidence among mothers residing in AL02-Qender and AL03-Jug regions compared to AL01-Veri is unlikely due to measured characteristics not included in the multivariable model, and is unclear. Regarding the association between parental education and vaccine coverage/confidence, the literature shows conflicting evidence [5,32,34,42,43].

### Limitations

Our study has limitations, and vaccine confidence was not analysed for individual vaccines. We considered immunisation timeliness as a proxy of vaccine confidence and other factors that may contribute to delaying or refusing childhood vaccines, like supply, access and health workforce, were not considered [26,44]. Childhood immunisation delay/refusal was based on maternal reporting, and as such, it is subject to information (recall) bias and poor memory recall, that may result in misclassification of the outcome. Poor memory recall or non-differential misclassification could lead to under-estimation of the observed associations. Differential misclassification would distort the strength of the associations also towards the null since mothers who do not adhere to immunisation schedules might feel uncomfortable with questions on vaccines and might not provide accurate answers due to social desirability bias. The work status of parents during the last 12 months was a proxy measure of their work at the time of childhood immunisation, which may result in residual confounding.

### Future directions

Our recommendations for further research include prospective design, retrieving immunisation data for individual vaccines from registries, defining a time-frame for vaccination timeliness, and a larger sample size to better identify the factors for declining immunisation timeliness and coverage among Albanian children.

## Conclusions

In conclusion, our analysis of the demographic and health survey indicates an inverse relationship between the Internet/social media as the main health information source and vaccine confidence, and persistence of inequities in vaccine confidence with regard to maternal education, maternal work status, and place of residence (region) in Albania. Thereby, the public health implications are that reinforcement of scientific evidence-based online communication about childhood immunisation, and monitoring anti-vaccination movements and vaccine hesitancy sentiments on Albanian Internet/social media would be beneficial. Since parents tend to search online for information that would confirm their original beliefs [45], traditional ways of promoting vaccination by healthcare professionals who enjoy confidence still remain important and enable targeting the inequities.

## Data Availability

Stata code is accessible at GitHub.
The 2017-2018 Albania DHS dataset is accessible upon registration on the DHS website.

https://github.com/d-mayerova/Vaccine_confidence_Albania

https://dhsprogram.com/data/dataset/Albania_Standard-DHS_2017.cfm

## Authorship contribution statement

DM and KA conceptualised the study and DM undertook the analysis and wrote the initial draft. KA contributed to the review and interpretation of results, and the final drafting of the manuscript. All authors have approved the final version.

## Acknowledgements

KA is supported by the Vaccine Impact Modelling Consortium (OPP1157270).

